# Co-administration of treatment for rifampicin-resistant tuberculosis and chronic hepatitis C virus infection: a TBnet and ESGMYC study

**DOI:** 10.1101/2021.10.16.21265043

**Authors:** Simone Tunesi, Damien Le Dû, Gina Gualano, Joan-Pau Millet, Aliaksandr Skrahin, Graham Bothamley, Xavier Casas, Delia Goletti, Christoph Lange, Maria Musso, Fabrizio Palmieri, Valérie Pourcher, Christophe Rioux, Alena Skrahina, Nicolas Veziris, Dzmitry Viatushka, Mathilde Jachym-Fréchet, Lorenzo Guglielmetti, for the TBnet, the ESGMYC, the French MDR-TB Group

**Author notes:** **Corresponding author:** Lorenzo Guglielmetti., Laboratoire de Bactériologie-Hygiène, Faculté de Médecine Sorbonne Université, 91 Boulevard de l’hôpital, 75634 Paris Cedex 13, France. The authors contributed equally to this study (Gina Gualano, Joan-Pau Millet, Aliaksandr Skrahin). ESCMID (European Society on Clinical Microbiology and Infectious Diseases) Study Group on Mycobacterial Infections. Members of the French MDR-TB Group are the following: Dhiba Marigot-Outtandy (Bligny), Xavier Lescure, Marie Dubert, and Yazdan Yazdanpanah (Paris-Bichat), Eric Caumes, Pascaline Choinier and Elie Haddad (Paris-Pitié-Salpetrière), Jakub Kowalczyk (Mulhouse), Hélène Laurichesse and Olivier Lesens (Clermont Ferrand), Alexandra Aubry, Isabelle Bonnet, and Florence Morel (Paris-CNR Mycobactéries). **Author contributions:** LG and ST made a substantial contribution to the conception and design of the work, to the acquisition, analysis and interpretation of data for the work, performed statistical analysis, wrote the manuscript, critically revised the manuscript for important intellectual content, gave final approval of the current version to be published, and agree to be accountable for all aspects of the work in ensuring that questions related to the accuracy or integrity of any part of the work are appropriately investigated and resolved. DLD and MJF made a substantial contribution to the conception and design of the work, to the acquisition and interpretation of data for the work, critically revised the manuscript for important intellectual content, gave final approval of the current version to be published, and agree to be accountable for all aspects of the work in ensuring that questions related to the accuracy or integrity of any part of the work are appropriately investigated and resolved. GG, JPM, and AS made a contribution to the conception and design of the work, to the acquisition and interpretation of data for the work, critically revised the manuscript for important intellectual content, gave final approval of the current version to be published, and agree to be accountable for all aspects of the work in ensuring that questions related to the accuracy or integrity of any part of the work are appropriately investigated and resolved. All other authors made a substantial contribution to the conception and design of the work, to the acquisition and interpretation of data for the work, critically revised the manuscript for important intellectual content, gave final approval of the current version to be published, and agree to be accountable for all aspects of the work in ensuring that questions related to the accuracy or integrity of any part of the work are appropriately investigated and resolved. **Financial and competing interest disclosure:** No funding to declare for the present work.

## Abstract

Concomitant treatment for chronic hepatitis C virus infection and multidrug-resistant tuberculosis is safe and effective.

## TEXT

Chronic hepatitis C virus (HCV)-infection affects worldwide 71 million people.[1] Direct-acting antivirals (DAA) revolutionised HCV clinical management since their introduction.[2,3] Tuberculosis is responsible of 1.4 estimated million deaths per year and multidrug-resistant/rifampicin-resistant tuberculosis (MDR/RR-TB) is a major public health issue worldwide.[4] Chronic HCV-infection is estimated at 7% among active tuberculosis patients,[5] reaching up to 30% in some settings among MDR/RR-TB.[6] This is clinically relevant, as HCV-infection is associated with higher rates of liver-related toxicity during anti-tuberculosis treatment,[7] and toxicity during treatment is ultimately linked to worse outcomes.[8] Currently, the World Health Organization (WHO) recommends treating all HCV patients above 12 years with pan-genotypic DAA.[9] Concomitant HCV and rifampicin-susceptible tuberculosis treatment is contraindicated due to drug-drug interactions. Conversely, no interactions are expected between second-line anti-tuberculosis drugs and DAA.[10] However, only limited evidence is available on the co-administration of these drugs.[11, 12] Therefore, the WHO makes no firm recommendation on concomitant MDR/RR-TB and HCV-treatment.[9] In our study, we aimed to assess the safety and effectiveness of concomitant treatment of chronic HCV-infection and MDR-/RR-TB.

We performed an observational cohort study across centres affiliated to the Tuberculosis Network European Trialsgroup (TBnet),[13] and the Study Group on Mycobacteria of the European Society of Clinical Microbiology and Infectious Diseases (ESGMYC). Consecutive patients with a confirmed diagnosis of MDR/RR-TB (phenotypic drug susceptibility testing) and HCV-infection (positive HCV polymerase chain reaction [PCR]) who started DAA during or up to four weeks before MDR/RR-TB treatment since January 1, 2015, were included. Patient follow-up was censored on February 15, 2021. Primary endpoints were sustained virologic response at 12 weeks and 24 weeks after finishing HCV-treatment, MDR/RR-TB treatment outcome,[14] rates of Grade 3 or higher liver-related adverse events, and total rates of serious adverse events (SAE). SAE were defined as events which were life-threatening or resulted in permanent disability, prolonged hospitalization, or death. De-identified data were collected retrospectively and collated in a secured database at the coordinating centre. Continuous variables were described using median with interquartile range (IQR), while frequency and proportions were reported for categorical variables. Statistical analysis was performed using STATA 12.0 (StataCorp, USA). Ethical approval was provided by the Institutional Review Board of the coordinating centre (Bligny Hospital, Briis-sous-Forges, France) and by other participating centres.

Overall, 23 patients were enrolled across six centres and four countries: three in France, and one in Belarus, Italy, and Spain. Two patients in the cohort were previously described.[12] Twenty were men (87%) and the median age was 42 years (IQR 39-45). Active smoking, alcohol abuse, and intravenous drug use were reported in 87%, 57%, and 35% of patients, respectively; 39% of patients were homeless. Nine patients were HIV-infected, with a median CD4 lymphocyte count of 85 cells/mm^3^ (IQR 77-626); among four for whom HIV-treatment status was known, three were receiving antiretrovirals (tenofovir disoproxil fumarate/emtricitabine plus darunavir [n=2] or raltegravir [n=1]) and one was not treated. One patient had HBV/HDV chronic hepatitis. Median baseline body mass index and serum albumin were 20 kg/m^2^ (IQR 18-21) and 38 mg/dl (IQR 34-42).

Among 20 patients with available HCV genotype, the predominant was 3 (40%), then 1a (30%), 1b (25%), and 4 (5%). Liver fibrosis, evaluated mainly by fibroscan and hepatic ultrasound, was absent (F0) or mild (F1) in the majority of patients (73%), and moderate (F2) in 18%. Two patients (9%) had liver cirrhosis; none had hepatocellular carcinoma. The most frequently used DAA were velpatasvir/sofosbuvir (39%) and sofosbuvir/daclatasvir (35%). Median DAA treatment duration was 84 days (IQR 83-91). The decision to start DAA was mostly due to previous hepatotoxicity during MDR/RR-TB treatment (30%) or because of elevated transaminases before starting MDR/RR-TB treatment (26%). DAA were usually started during MDR/RR-TB treatment (65%), a median of 267 days (IQR 69-584) after MDR/RR-TB treatment start. All patients completed DAA treatment without interruptions. At baseline, plasmatic HCV-RNA was detected in all patients, with median levels of 5,7 log IU/ml (IQR 5.2-6.3) among 13 patients who performed quantitative evaluation. Plasmatic HCV-RNA detection decreased to 31% of patients at week 4 and 8 of treatment and became undetectable for all patients starting from week 12 (Figure 1). Assessment of sustained virological response at week 4, 12, and 24 after the end of HCV-treatment was available for 11 patients and HCV-RNA was undetectable in all.

**Figure 1.**
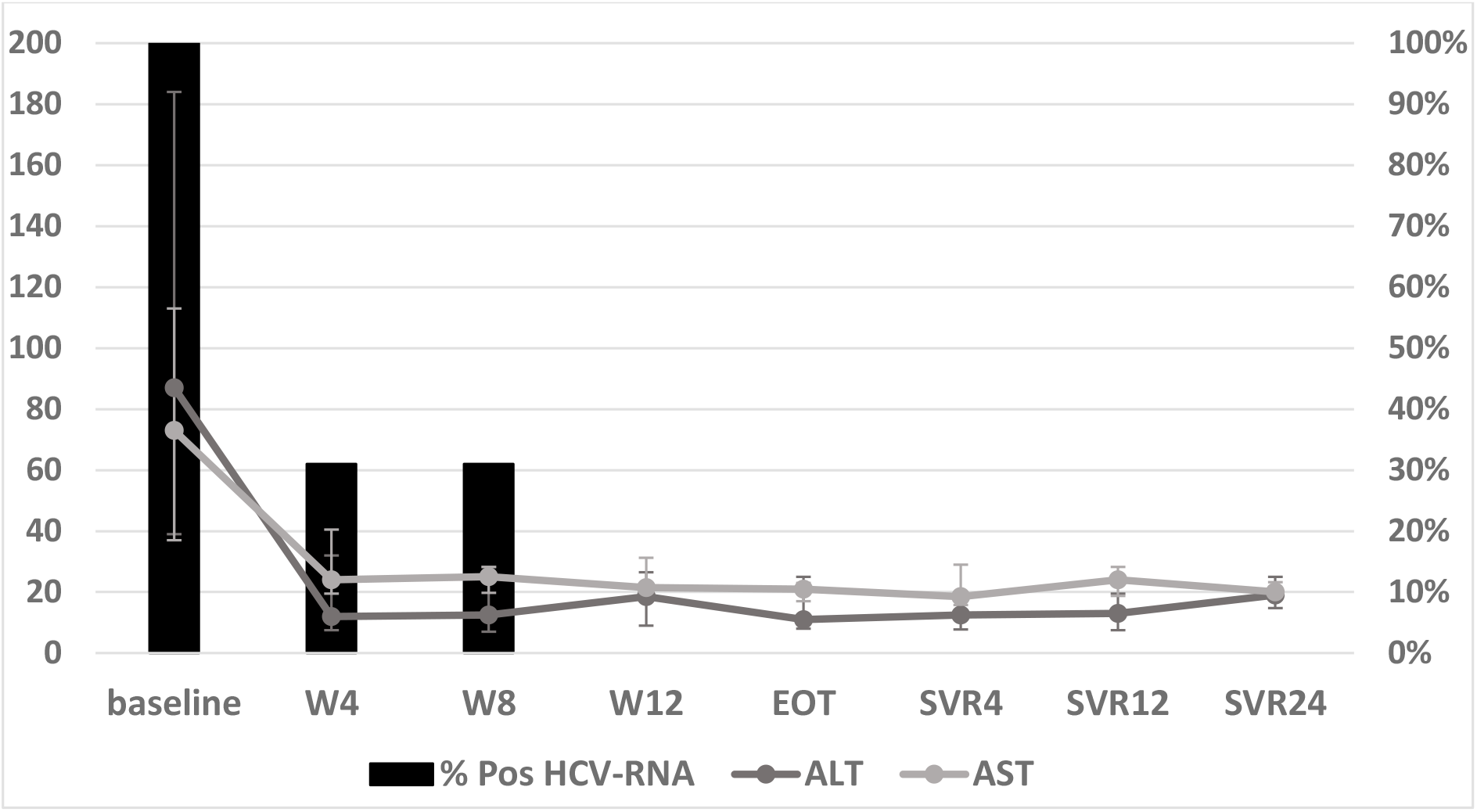
Plasma HCV-RNA positivity rates (%) and blood transaminase levels (U/L, with standard deviations bars) during treatment with direct-acting antivirals in a cohort of 23 HCV-infected patients with active rifampicin-resistant tuberculosis. W4, W18, W12 = 4, 8, and 12 weeks after starting HCV treatment; EOT = at end of HCV treatment; SVR4, SVR12, SVR24 = 4, 12, and 24 weeks after the end of HCV treatment.

All patients had pulmonary tuberculosis, with associated-extrapulmonary involvement in 4 (17%). Bilateral lung involvement and lung cavitations were present in 61% and 46% of patients, respectively. Sputum smear was positive at baseline for 50%. Overall, 52% of patients had received previous tuberculosis treatment, 30% with first-line and 22% with second-line drugs. According to phenotypic drug-susceptibility testing, 53% had MDR/RR-TB with no additional resistance to fluoroquinolone and second-line injectables, 30% had additional resistance to one of these two drug classes, and 17% to both. All patients received linezolid, and most received clofazimine (87%), cycloserine (78%), and bedaquiline (65%). At censoring, 52% of patients were still ongoing MDR/RR-TB treatment. Among the other 11 patients, 10 (91%) achieved cure and one (9%) died of accidental causes.

Overall, 18 liver-related adverse events were reported across 11 patients (48%): the majority (94%) occurred during MDR/RR-TB treatment but before DAA were started. Most adverse events were Grade 1 (56), while 22% were Grade 2 and 3, respectively. No grade 4 adverse events or liver-related SAE were reported. In 30%, the adverse event led to the temporary or permanent discontinuation of an anti-tuberculosis drug. Blood transaminase were increased at the beginning of DAA treatment (AST: median 60 U/l [IQR 37-102], ALT: median 79 U/l [IQR 37-167]) but decreased into normal range from week 4 of treatment (Figure). The median duration until resolution was 90 days (IQR 50-147). Other non-liver-related SAE were observed in seven patients (30%), mostly linked to peripheral neuropathy (n=3), anaemia and hearing loss (n=2, each).

In our multicentre, retrospective cohort study, we found that concomitant HCV and MDR/RR-TB treatment was effective and well-tolerated. DAA treatment led to achieve undetectable HCV-RNA and sustained virological response for all patients with available data. Similarly, treatment success was achieved for 91% of patients who completed MDR/RR-TB treatment. These results are particularly important considering the high rates of HIV infection, as well as the high prevalence of HCV genotype 3. Similar, encouraging outcomes have been reported in a cohort from Armenia.[11] Liver-related adverse events were mostly mild or moderate, and occurred mainly before the start of DAA. Moreover, blood transaminases decreased into normal range for all patients from week 4 of DAA treatment. Considering that HCV-treatment was often started because of previous hepatotoxicity or increased transaminases, our results suggest that co-administration of DAA may prevent liver toxicity during MDR/RR-TB treatment.

Our study is limited by the small sample, retrospective data collection, and lack of MDR/RR-TB outcome in patients whose treatment is still ongoing. However, the results are relevant, as we show that the association of second-line anti-tuberculosis drugs and DAA seems safe, without impact of drug-drug interactions on outcomes. Treatment with DAA should be considered in MDR/RR-TB patients to reduce tuberculosis-treatment-related hepatotoxicity, minimizing the risk of prolonged treatment interruption, and prevent progression of HCV-mediated liver disease. Integrated services for the management of tuberculosis, HIV, and HCV, should be promoted widely and collaboration with surveillance, prevention and control programs should be strengthened.[15]

## Data Availability

All data produced in the present study are available upon reasonable request to the authors

## Notes

### Competing Interest Statement

The authors have declared no competing interest.

### Funding Statement

This study did not receive any funding

### Author Declarations

Ethical approval was provided by the Institutional Review Board of the coordinating centre (Bligny Hospital, Briis-sous-Forges, France) and by other participating centres.

